# Potential for High Dynamic Range Sedia Limiting Antigen Antibody Assay to Support Viral Load monitoring during Antiretroviral Therapy

**DOI:** 10.1101/2023.01.26.23285058

**Authors:** Joseph B. Sempa, Alex Welte

## Abstract

**Introduction:** Viral Load (VL) monitoring is a crucial component of patient care during antiretroviral therapy (ART) but is not routinely available in many resource-constrained settings, where millions of patients will require care for decades to come. We hypothesise a serologic ‘recent infection’ test (Sedia LAg assay) which has a high dynamic range for detecting antigen-driven antibody response can provide informative proxies for VL trajectories.

**Methods:** We analysed data linked via specimens in a well-described repository for recent infection test benchmarking (CEPHIA collaboration). Patient panels were comprised of 1) observations straddling ART start; 2) observations from a period of stable viral suppression; 3) observations straddling rebound after a period of viral suppression. We analysed an individual’s Sedia LAg ELISA normalised optical density (ODn) trends within these categories. Using groups 2) and 3) we evaluated the specificity and sensitivity of a proposed proxy for “the latest observation is at a time of VL rebound”; proxy was defined as follows: we estimated patient-specific mean-previous-ODn for all observations with at least two preceding virally suppressed observations. We considered various thresholds to define both “VL suppression” and “ODn uptick”.

**Results:** In regression analysis by category: 1) ODn gradients are statistically significantly negative just after ART-start (p=0.010); 2) During periods of stable viral suppression, ODn tended to decline, but not statistically significantly, for a range of clinically meaningful “VL suppression” thresholds; 3) comparing ODn values just before, versus at, “VL rebound”, ODn changes were statistically significantly increasing at rebound (p= 0.001). In the analysis comparing groups 2) and 3), at a Z score threshold of 0.8, the proposed proxy for a first viral rebound had an observed specificity and sensitivity both close to 90%.

**Conclusion:** The high dynamic range of serological tests previously investigated for defining ‘recent infection’ has potential, as demonstrated using the Sedia LAg ELISA, to provide meaningful information about the success of ART, during treatment initiation, at times of stable suppression, and to flag possible viral rebound. It should be investigated how this can be combined with patient management workflows and (clinical and) other data, to provide efficiencies in long-term monitoring viral control in resource-limited settings.

## Introduction

Patient monitoring of ART is a long-term challenge requiring efficient use of resources and maximally informative use of biomarkers. In particular, detecting suboptimal treatment, and then efficiently and correctly identifying whether mal-adherence or resistance is the main underlying cause, is a matter of great health and financial impact. It will also become increasingly important to relieve pressure on the drugs pipeline, in which investment is expected to ultimately decline as the epidemic (hopefully) cools off over the next generation or two. Currently, viral load (VL) monitoring is expensive and not universal, it requires dedicated platforms, and it provides a narrow time window of clinically relevant information. In highly functional routine care settings outside of studies, plasma viral load is assessed about once a year [1]. There may be significant practical value in more time-averaged markers of viral replication and suppression, such as immune system response markers which naturally respond gradually to pathogen levels, and which are measurable using routine diagnostic systems. Conceptually, this is analogous to the use of the HbA1C marker, which offers a more time-averaged indicator of blood sugar levels, for the management of diabetes, than does a single time point insulin level measurement [2].

Selleri et al. 2007 showed that antibody trajectory keeps track of the viral load after ART start even in periods of treatment interruptions, however, they had data on only 4 subjects [3]. Further, Keating et al 2017, using assays developed for infection staging in the sense of identifying ‘recent’ infections for surveillance, showed declining HIV antibodies during ART in early or late-treated patients and Elite Controllers [4]. However, these patients were sampled at specific time points straddling ART initiation and all these patients were suppressed after ART. The behaviour of HIV antibodies and viral load response trajectories after ART initiation across different groups of viral suppression is still unknown.

Serological diagnostic assays are the most fundamental component of any HIV treatment programme. Data from the use of such assays, including so-called ‘recency’ or more broadly ‘infection staging’ assays in routine testing is increasingly being used for both post-surveillance purposes and post-test counselling, but to date, we are not aware that immuno-assays have been systematically investigated for utility in routine monitoring of treatment stability.

Beyond patient management, there is broad interest in better understanding cumulative exposure to viremia, and how this affects mortality, classic opportunistic infections as well as nominally non-AIDS-related lymphomas, and immune system recovery - and we hope the present analysis can help develop useful proxies for cumulative viremic exposure.

## Methods

We analysed data from the Consortium for the Evaluation and Performance of HIV Incidence Assays (CEPHIA) database. The CEPHIA ‘Evaluation Panel’ [5] has 2424 total samples tested using the Sedia LAg assay, from 928 unique specimens of patient-study interactions, with a wide range of times since infection, mostly infected with HIV-1 subtype B (25.6%), C (42.6%), A1 (20%), and D (8.8%). For this analysis, we used the longitudinal data for 233 unique patients, whom we grouped into three categories, 1) observations straddling ART start (n = 18); 2) observations from a period of stable viral suppression (n = 95); 3) observations straddling rebound after a period of viral suppression (n = 120).

### Patient and Specimen Data

Laboratory testing was done by the CEPHIA collaboration as part of an early multi-assay evaluation for the suitability of various assays to be used for incidence estimation. Both the clinical background information and the LAg assay results which were used in this study were accessed from a secondary analysis [6] which provided open access to this data.

### Statistical analysis

For the purpose of the present analysis, we define a *suppressed* viral load as one that is less than 1000 copies/mL. We considered available data from three groups of patients:

1. Patients who were observed both before and after initiation of ART
2. Patients who maintained a suppressed viral load at all times under observation
3. Patients who were observed with an initially suppressed viral load, and then were observed to have an unsuppressed viral load at a later timepoint(s)

For each individual patient trajectory in category one and category two, we estimated the slope of the LAg normalized optical density (ODn) curve and tested the null hypothesis that this slope is zero. We also estimated the differences in successive Sedia LAg ODn readings, without regard to time intervals, and tested the null hypothesis that successive Sedia LAg results are from the same distribution, without drift.

For category three, we observed the change in LAg ODn between

- the last virally suppressed visit and the first virally unsuppressed visit (when viral load became unsuppressed after a period of at least two suppressed viral loads)
- a (latest) virally unsuppressed visit, and a subsequent virally suppressed visit (when an unsuppressed viral load was followed by a suppressed viral load)

In both cases, we tested the hypothesis that such successive measurements differ only by measurement noise. To estimate the magnitude of inter-measurement fluctuations, which is needed to evaluate the significance of observed differences, we used all observations of the subjects in group 2, and obtained the classic ‘pooled variance’ estimate.

In patients who are only ever seen as virally suppressed and have at least two visits, we performed two variations on the theme of tests of the null hypothesis that there is no drift in ODn readings.

1. we compared each Sedia LAg value with the mean Sedia LAg values of all *previous* Sedia LAg readings.
2. we compared each Sedia LAg result with the average of *all* the other readings the patient had.

In both cases, the hypothesis was that the current Sedia LAg value is the same as the mean Sedia LAg value if the patient is virally suppressed.

Among those who were experiencing loss of viral suppression with at least two suppressed visits before they were virally unsuppressed, we compared the Seadi LAg value when they became unsuppressed to the mean Sedia LAg value for the prior visits.

Using the pooled variance estimate from subjects who were always viral suppressed, we assigned each observation of LAg ODn on the date of the first unsuppressed viral load a Z score – simply in the sense of expressing the LAg reading as a multiple of standard deviations from the subject-specific estimated mean, not to imply that these Z scores should be expected to be normally distributed. We are aware (and our results show) that the distribution of ODn values, both within and between individuals, is NOT normal. Using this ‘Z score’, and a range of thresholds, we define ‘ODn uptick’ simply as an ODn reading that has a ‘Z score’ greater than the candidate threshold. We then estimate the sensitivity and specificity of ‘ODn uptick’ as a marker for ‘loss of viral suppression’.

All statistical analyses were carried out using R, version 4.0.2 (R Foundation for Statistical Computing, Vienna, Austria)

## Results

Figure 1 shows the distribution of slopes obtained by fitting simple linear (time) regression models to the LAg assay ODn values of those individuals who were at all times virally suppressed. The average of the individual regression slopes among these patients was −1.93×10^−5^ units of ODn per day, which was not statistically significantly different from zero (P > 0.9). Individual model slopes can clearly be seen to cluster around zero. According to the individual regressions, all but one of the subjects’ Sedia LAg slope results were indistinguishable from zero (P values > 0.05), which is very close to the expected number of accidentally ‘significant’ slopes.

**Figure 1:**
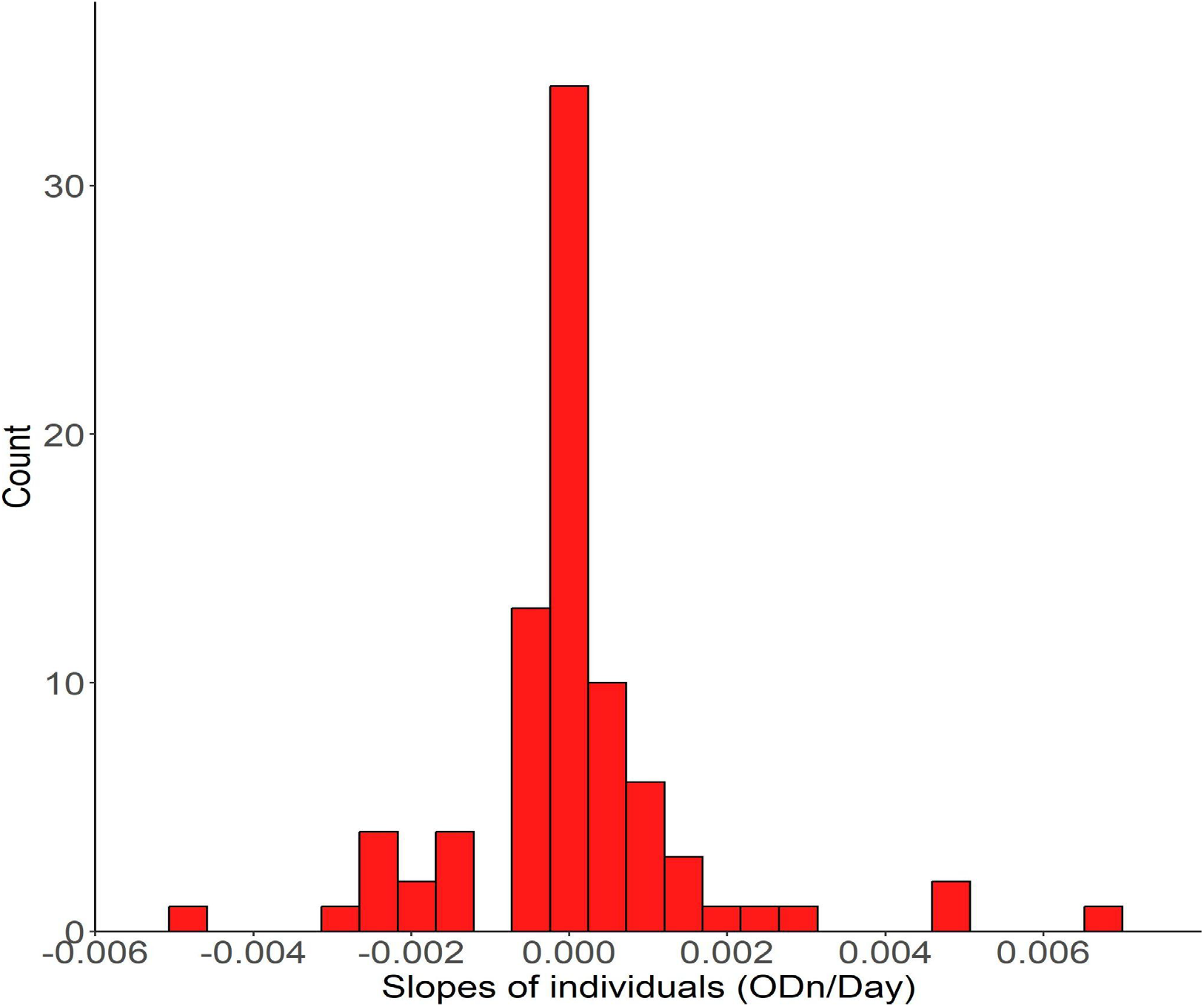
Frequency distribution of fitted slopes of Sedia LAg ODn values among patients who were ‘virally suppressed’ (Viral load below 1000 copies/ml) at all times while under observation.

In figure 2, we present distributions of differences in LAg ODn from successive observations under various scenarios. Panel A shows all successive time point pairs from subjects who were virally suppressed at all times while under observation. As would be expected from the regression analysis, there is no obvious evidence that ODn values drift in any significant way as time passes. Panel B shows the distribution of LAg ODn differences for time point pairs in which the first time point is the last of at least two visits in which viral load was suppressed, before becoming unsuppressed – and hence the second time point is the date on which the viral load was first seen to have become unsuppressed. The distribution looks visibly biased towards upticks in ODn values associated with the upticks in viral load. Panel C shows the opposite process from panel B – in the sense that we show the changes in ODn which accompany a change in VL from above to below our threshold of ‘suppression’.

**Figure 2:**
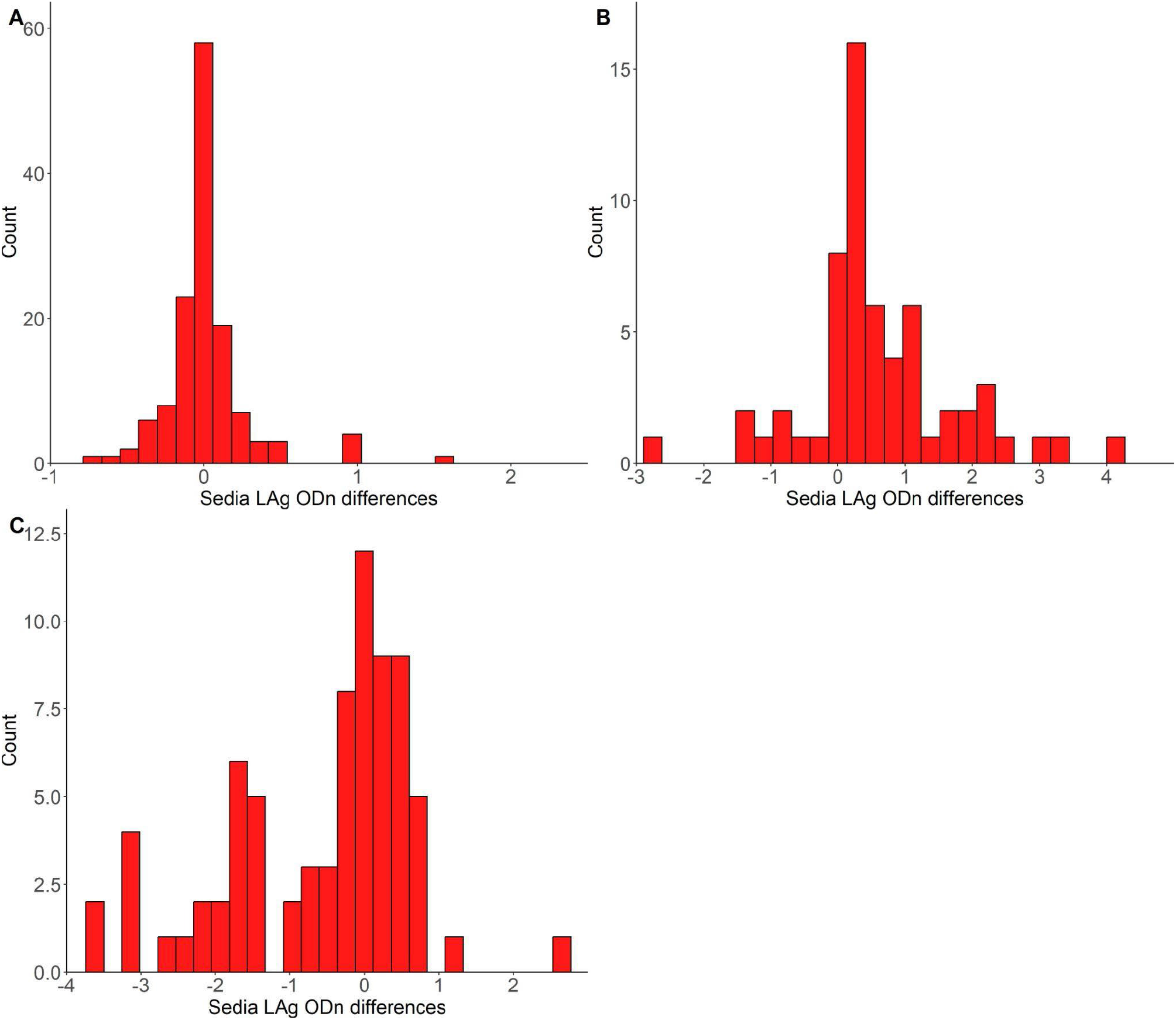
Frequency distribution of the Sedia LAg ODn differences between successive visits for various scenarios. **A -** All successive pairs of time points from patients who were virally suppressed (< 1000 copies/mL) throughout follow-up; **B -** Change in ODn from ‘last’ virally suppressed visit to ‘first’ visit where VL was not suppressed. **C -** Change in ODn from ‘last’ virally unsuppressed visit to ‘first’ visit where VL was suppressed.

A key point of the present analysis is to see whether it is possible to find a pattern of LAg ODn values which might be a proxy for meaningful patterns in the patient viral load. Inspired by figure 2, we proposed, as described in the methods, assigning patient ODn values a ‘Z score’ by comparing it to previous ODn values of that patient and normalizing, for the present purposes, by the pooled variance estimate from all subjects who were always virally suppressed. In figure 3 we plot the distribution of nominal ‘p-values’ implied by these Z scores. In panel A, we see the p-value distribution from the point of view of following patients up in real-time, and at each visit comparing the latest ODn value to the mean of all the previous ones for that patient.

**Figure 3:**
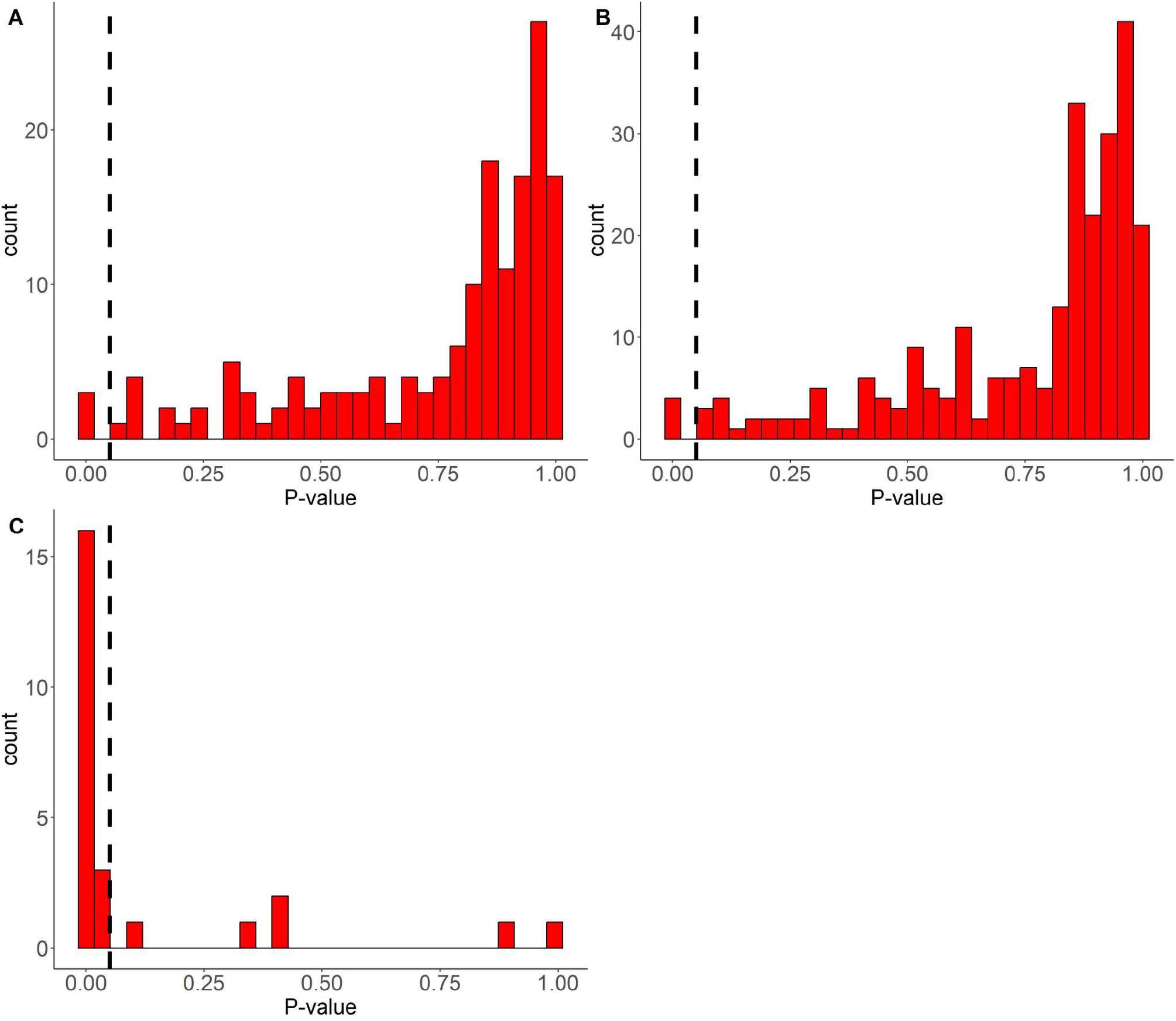
Frequency Distribution of p-values for proposed test for ‘ODn Uptick’. **A -** When comparing the current Sedia LAg value with the average of previous values, of a patient who was virally suppressed throughout follow-up; **B -** When comparing any Sedia LAg value with the mean of all other values ever obtained from a patient who was virally suppressed throughout follow-up; **C -** When comparing the Sedia LAg value of a patient, at the time point when VL >1000 copies/ml for the first time after a period of viral suppression, with the mean of the Sedia LAg values from their previous visits when VL was suppressed.

For comparison, we show in panel B what the distribution of nominal p-values would be if we compare the ODn value from each visit to all the other ODn values from all other values at hand for that patient – even if they are from later times. While this is not possible to evaluate in real-time monitoring, it leads to essentially the same distribution of nominal values, which indicates that it is meaningful to talk of a natural distribution from which patients’ Z scores are drawn. However, the fact that this distribution of ‘p values’ is decidedly different from uniform – in particular, substantially enriched for values close to 1, shows that the underlying ODn values cannot be very well approximated as coming from a normal distribution. We do not pursue this further at this point, given the limitations of our data and our initial aspirations.

In panel C, we observe the distribution of nominal p values of our ‘Z scores’ as observed at time points where patients exhibit the first ‘viral rebound’ (first unsuppressed viral load after at least two suppressed viral loads). It is striking that they are clustered at low values, suggesting there may be scope for a statistically meaningful signal in this proposed Z statistic. Recalling, as just noted, that the nominal p-value distribution is clearly indicative of a non-normal underlying distribution, we should not over-interpret the p-values in themselves.

Reverting to the basic Z scores which can be assigned to patients at each visit from the third visit, we can now consider, without leaning on the interpretation of this into a formal p-value, simply defining a threshold on the Z statistic, and thus defining the notion of ‘ODn uptick’. By considering the Z values obtained throughout our data set, at times when there was

1. a pattern of prior viral load suppression, and
2. a directly observed transition in VL from suppressed to unsuppressed (or not)

we can define an observed sensitivity and specificity of this ‘ODn Uptick’ as a proxy for ‘End of viral load suppression’. In figure 4, we plot, without considering uncertainties in this preliminary analysis, this sensitivity and specificity. Note that at Z score thresholds from about 0.5 to 1.0, both sensitivity and specificity are over 80%, which suggests significant information about VL trajectories is being reflected in the LAg ODn trajectories.

**Figure 4:**
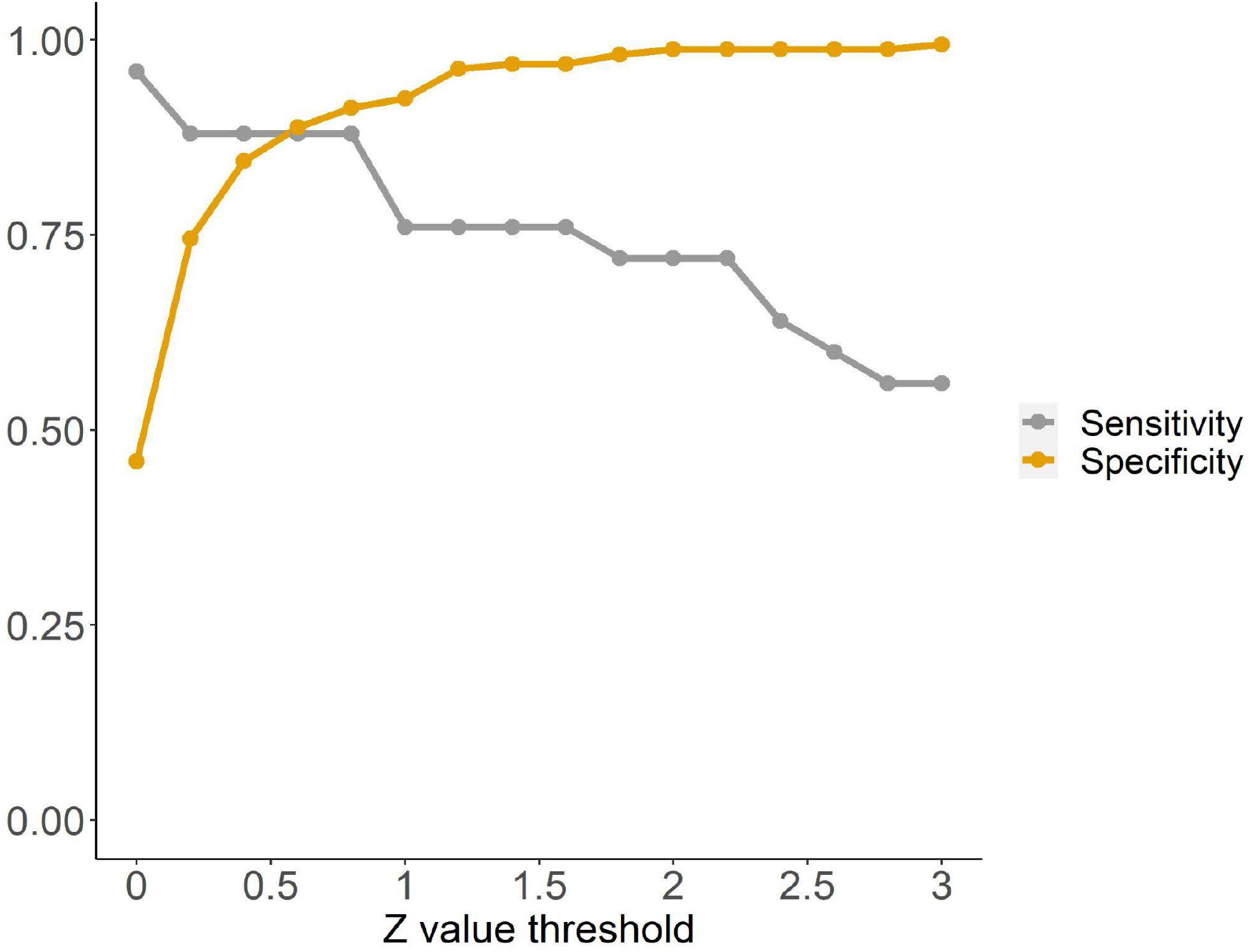
Sensitivity and Specificity for detecting an initial viral resurgence, as a function of ‘Z-value threshold’ in comparing a most recent Sedia LAg ODn to the mean of preceding values obtained during viral suppression.

## Discussion

To our knowledge, this is the first analysis that directly looks at the relationship between

- clinically meaningful changes in HIV viral load levels during antiretroviral therapy, and
- markers obtainable by applying high dynamic range antibody assays

Related, but less directly treatment-monitoring-focused, investigations of the relationship between high dynamic range serology and viral load have previously been conducted, for example by Selleri et al. 2007 [3] and Keating et al. 2017 [4]. Our findings support these previous analyses, and suggest, even more pointedly, that the Sedia LAg assay (better known as a frontrunner in this class of assays developed for the primary purpose of HIV surveillance by providing efficient estimates of HIV incidence through the creation of a category of nominally ‘recent infections’) contains significant useful information along these lines. In particular

- Patients stable on antiretroviral therapy show no statistically detectable (at our resolution thus far) drift in LAg ODn values over time
- A majority of patients experiencing viral load rebound show a statistically detectable uptick in the LAg ODn readings
- Patients on a trajectory of attaining viral load suppression show statistically weak tendencies of having declining values of LAg ODn.

This seems to us a very promising initial finding along these lines and suggests that several additional questions should be investigated.

Some are of a detailed statistical nature, such as the prospects for changing from a population-level pooled variance estimate of ODn fluctuations to a patient-specific variance estimate, which might significantly improve monitoring over the long-time frames associated with ART monitoring.

Other more complex ‘healthcare system’ level questions probe how it might be feasible to achieve goals like

- Monitor patients early after ART initiation to support distinguishing those who are responding best from those who are responding least well
- transition patients into serology-based monitoring after it has been more conventionally established that initial treatment has led to viral suppression.

We will continue to work with various partners to produce or access additional appropriate data linking high dynamic range serology to viral load measurements, and we also hope that others, who are working in the more overarching health system dynamics, will entertain further investigations into how this statistical evidence can be meaningfully used to make treatment monitoring more efficient or more effective, or both.

## Data Availability

All data produced are available online at https://doi.org/10.1371/journal.pone.0220345.s008

https://zenodo.org/record/4900634#.Y9Kcl3ZByUk

## Acknowledgements

The Dr Joseph B Sempa received funding from the European and Developing Countries Clinical Trials Partnership (EDCTP) under the Career Development Fellowship TMA2019CDF-2760. He also acknowledges the contribution of Prof, Christopher Viljoen as his mentor on this project.

This work is based on research supported by the (South African) Department of Science and Innovation and the National Research Foundation. Any opinion, finding, conclusion or recommendation expressed in this material is that of the authors and the NRF does not accept any liability in this regard.

The Consortium for the Evaluation and Performance of HIV Incidence Assays (CEPHIA) comprises: Alex Welte, Joseph Sempa, formerly: David Matten, Hilmarie’ Brand, Trust Chibawara (South African Centre for Epidemiological Modelling and Analysis, Stellenbosch University); Gary Murphy, Jake Hall, formerly: Elaine Mckinney (Public Health England); Michael P. Busch, Eduard Grebe, Shelley Facente, Dylan Hampton, Sheila Keating, formerly: Mila Lebedeva (Vitalant Research Institute, formerly Blood Systems Research Institute); Christopher D. Pilcher, Kara Marson (University of California San Francisco); Reshma Kassanjee (University of Cape Town); Oliver Laeyendecker, Thomas Quinn, David Burns (National Institutes of Health); Susan Little (University of California San Diego); Anita Sands (World Health Organization); Tim Hallett (Imperial College London); Sherry Michele Owen, Bharat Parekh, Connie Sexton (Centers for Disease Control and Prevention); Matthew Price, Anatoli Kamali (International AIDS Vaccine Initiative); Lisa Loeb (The Options Study—University of California San Francisco); Jeffrey Martin, Steven G Deeks, Rebecca Hoh (The SCOPE Study—University of California San Francisco); Zelinda Bartolomei, Natalia Cerqueira (The AMPLIAR Cohort— University of São Paulo); Breno Santos, Kellin Zabtoski, Rita de Cassia Alves Lira (The AMPLIAR Cohort—Grupo Hospital Conceição); Rosa Dea Sperhacke, Leonardo R Motta, Machline Paganella (The AMPLIAR Cohort—Universidade Caxias Do Sul); Esper Kallas, Helena Tomiyama, Claudia Tomiyama, Priscilla Costa, Maria A Nunes, Gisele Reis, Mariana MSauer, Natalia Cerqueira, Zelinda Nakagawa, Lilian Ferrari, Ana P Amaral, Karine Milani (The São Paulo Cohort—University of São Paulo, Brazil); Salim S Abdool Karim, Quarraisha Abdool Karim, Thumbi Ndungu, Nelisile Majola, Natasha Samsunder (CAPRISA, University of Kwazulu-Natal); Denise Naniche (The GAMA Study—Barcelona Centre for International Health Research); Inácio Mandomando, Eusebio V Macete (The GAMA Study—Fundacao Manhica); Jorge Sanchez, Javier Lama (SABES Cohort— Asociación Civil Impacta Salud y Educación (IMPACTA)); Ann Duerr (The Fred Hutchinson Cancer Research Center); Maria R Capobianchi (National Institute for Infectious Diseases “L. Spallanzani”, Rome); Barbara Suligoi (Istituto Superiore di Sanità, Rome); Susan Stramer (American Red Cross); Phillip Williamson (Creative Testing Solutions / Vitalant Research Institute); Marion Vermeulen (South African National Blood Service); and Ester Sabino (Hemocentro do São Paolo). For enquiries, please contact Gary Murphy (Public Health England) at.

## Author contributions

**Conceptualization:** Joseph B. Sempa, Alex Welte

**Data curation:** Joseph B. Sempa

**Formal analysis:** Joseph B. Sempa, Alex Welte.

**Funding acquisition:** Joseph B. Sempa, Alex Welte.

**Methodology:** Joseph B. Sempa, Alex Welte.

**Project administration:** Joseph B. Sempa.

**Resources:** Joseph B. Sempa

**Software:** Joseph B. Sempa, Alex Welte.

**Supervision:** Alex Welte.

**Visualization:** Joseph B. Sempa, Alex Welte.

**Writing – original draft:** Joseph B. Sempa, Alex Welte.

**Writing – review & editing:** Joseph B. Sempa, Alex Welte

## References

1 Lecher S, Williams J, Fonjungo PN, et al. Progress with Scale-Up of HIV Viral Load Monitoring - Seven Sub-Saharan African Countries, January 2015-June 2016. MMWR Morb Mortal Wkly Rep 2016;65:1332–5. doi:10.15585/mmwr.mm6547a2

2 Roche Diagnostics. cobas HbA1c Test 10. 2013. http://www.roche.com (accessed 22 Jan 2019).

3 Selleri M, Orchi N, Zaniratti MS, et al. Effective highly active antiretroviral therapy in patients with primary HIV-1 infection prevents the evolution of the avidity of HIV-1-specific antibodies. J Acquir Immune Defic Syndr 2007;46:145–50. doi:10.1097/QAI.0b013e318120039b

4 Keating SM, Pilcher CD, Jain V, et al. HIV Antibody Level as a Marker of HIV Persistence and Low-Level Viral Replication. Journal of Infectious Diseases 2017;216:72–81. doi:10.1093/infdis/jix225

5 Kassanjee R, Pilcher CD, Keating SM, et al. Independent assessment of candidate HIV incidence assays on specimens in the CEPHIA repository. AIDS 2014;28:2439–49. doi:10.1097/QAD.0000000000000429

6 Sempa JB, Welte A, Busch MP, et al. Performance comparison of the Maxim and Sedia Limiting Antigen Avidity assays for HIV incidence surveillance. PLoS One 2019;14:e0220345. doi:10.1371/journal.pone.0220345

